# Binocular integration of perceptually suppressed visual information in amblyopia

**DOI:** 10.1101/2020.06.04.20122440

**Authors:** Amy Chow, Andrew E. Silva, Katelyn Tsang, Gabriel Ng, Cindy Ho, Benjamin Thompson

## Abstract

Abnormal visual experience during an early critical period of visual cortex development can lead to a neurodevelopmental disorder of vision called amblyopia. A key feature of amblyopia is interocular suppression, whereby information from the amblyopic eye is blocked from conscious awareness when both eyes are open. Suppression of the amblyopic eye is thought to occur at an early stage of visual processing and to be absolute. Using a binocular rivalry paradigm, we demonstrate that suppressed visual information from the amblyopic eye remains available for binocular integration and can influence overall perception of stimuli. This finding reveals that suppressed visual information continues to be represented within the brain even when it is blocked from conscious awareness by chronic pathological suppression. These results have direct implications for the clinical management of amblyopia.

## Introduction

Normal development of the visual pathway is critically dependent on congruous binocular visual experience. Disruption of visual development by the misalignment of one eye (strabismus) or a difference in focusing power between the two eyes (anisometropia) can result in amblyopia, a neurodevelopmental disorder of vision that is characterized by both monocular and binocular visual impairments(1–3). Interocular suppression is a key component of amblyopia, whereby information from the affected amblyopic eye is blocked, or suppressed, from conscious awareness when both eyes are open. Suppression may occur to avoid visual confusion caused by conflicting input from the two eyes and it renders a structurally binocular visual system functionally monocular in individuals with amblyopia (4–6). It has been assumed that suppression of the amblyopic eye is absolute and visual information from the suppressed eye does not contribute to visual processing. As a result, adults with amblyopia are often not given refractive correction for their amblyopic eye because it is considered to be of no added benefit(7).

Interocular suppression can be induced in individuals with normal vision by presenting conflicting images to each eye. Perception alternates between the two dichoptically presented images as one eye is suppressed and the other becomes dominant (a phenomenon known as binocular rivalry). There is evidence that visual information from a suppressed eye continues to be available for neural processing despite being blocked from conscious awareness in individuals with normal vision. Specifically, dichoptic presentation of orthogonal moving gratings can induce the percept of a single grating moving in an integrated motion direction (8–10). This suggests that integration of motion information from both eyes can take place even when form information from one eye (the grating) is suppressed.

In this study we explored the possibility that suppressed visual information can influence perception in individuals with amblyopia, a disorder associated with chronic, pathological suppression of one eye due to abnormal visual cortex neurodevelopment. In particular, we used dichoptically presented moving gratings in a binocular rivalry paradigm to test for binocular integration of motion information in the presence of amblyopic eye suppression.

## Results

As expected, the dichoptic grating stimuli induced longer periods of non-dominant/amblyopic eye suppression in participants with amblyopia (n = 20; mean±SE: 19.9±2.3 minutes) than in participants with normal vision (n = 20; 8.97±0.7 minutes). Data were further analyzed only from participants who reported non-dominant/amblyopic eye suppression for at least 2 minutes (representing at least 5% of total viewing time, n = 16 participants with normal vision and 19 participants with amblyopia). Binocular integration of motion information from the suppressed non-dominant/amblyopic eye occurred 48±6% of the time in participants with normal vision and 31±6% of the time in participants with amblyopia (t_33_ = –1.995, p = 0.054). Notably, the proportion of time that participants with amblyopia experienced binocular motion integration under these conditions was significantly non-zero (t_18_ = 5.4, p<.00002).

## Discussion

Our results demonstrate that despite chronic and pathological suppression of the amblyopic eye, visual information suppressed from conscious awareness remains available for visual processing and can influence visual perception. The amblyopic visual system may operate similarly to the normal visual system where suppressed visual information retains a presence within the brain(11–13) and can influence perception (8–10, 14, 15). We conclude that suppression in amblyopia is not absolute and is likely to involve higher level brain areas that lie beyond the regions responsible for binocular motion integration. In addition, our observation of binocular motion integration in amblyopia supports the underlying theory behind binocular approaches to amblyopia treatment, which posits a structurally intact but functionally suppressed binocular visual system in amblyopia (4–6). Our results also have direct implications for clinical practice, suggesting that there may be value in providing appropriate optical correction for the amblyopic eye in adult patients not only for visual acuity improvement (7) but also for enhancing the quality of visual information available for binocular integration.

## Materials and Methods

Participants with normal vision (n = 20, mean±SE 23.9±0.7 years, 16 females) and amblyopia (n = 20, 39.5±3.1 years, 9 females) took part in this study. All participants were naïve to the experimental hypothesis and were reimbursed for their time. Participants provided written informed consent to take part in the study, and the study protocol was approved by the institutional ethics committee at the University of Waterloo, in accordance with the Declaration of Helsinki. Participants were screened at the School of Optometry and Vision Science at the University of Waterloo, Waterloo, ON or at the Mount Pleasant Optometry Centre, Vancouver, BC. Clinical assessment included visual acuity (using an electronic ETDRS chart), eye alignment (distance and near cover test) and stereoacuity (Randot Preschool Stereotest; Stereo Optical Co. Inc., Chicago, IL, USA). Amblyopia was defined as a minimum of a 2 logMAR line difference in visual acuity between the eyes, caused by either anisometropia (> 1 dioptre difference in spherical equivalent between the eyes or > 1.5 dioptres of cylinder in one eye) and/or strabismus (including history of strabismus surgery), with otherwise normal ocular and general health. All participants wore their habitual correction as needed. The dominant eye in participants with normal vision was determined as the eye more sensitive to blur as a +2.00 DS lens was held over one eye while binocularly observing letters 2 logMAR lines above their best-corrected visual acuity. The dominant eye in participants with amblyopia was defined by the eye with better best-corrected visual acuity. Clinical details for individuals with amblyopia are summarized in Table 1. Data and analysis code are available online in the University of Waterloo’s repository.

**Table 1.** Clinical details for participants with amblyopia. Some identifier codes are omitted as some participants could not fuse in the stereoscope. M = male, F = female, VA = visual acuity, A = anisometropia, S = strabismus, M = mixed (anisometropia and strabismus), NS = non-strabismic, XP = exophoria, RET = right esotropia, Δ = prism diopters.

| ID | Type | VA (FE) | VA (AE) | Stereoacuity | Ocular Deviation (Near) |
| --- | --- | --- | --- | --- | --- |
| A01 | A | 20/20 | 20/100 | >800" | NS, 12Δ XP |
| A02 | A | 20/20 | 20/40 | >400" | NS, ortho |
| A03 | A | 20/20 | 20/50 | >800" | NS, ortho |
| A04 | A | 20/25 | 20/200 | >800" | NS, ortho |
| A05 | M | 20/20 | 20/60 | >800" | 4Δ RET |
| A06 | M | 20/15 | 20/80 | >800" | 4Δ LXT |
| A07 | A | 20/20 | 20/25 | 60" | NS, ortho |
| A08 | A | 20/15 | 20/200 | >800" | NS, 2Δ XP |
| A09 | S | 20/15 | 20/25 | >800" | 17Δ LXT |
| A10 | A | 20/15 | 20/50 | >800" | NS, ortho |
| A11 | S | 20/15 | 20/30 | >800" | 8Δ RET |
| A12 | A | 20/15 | 20/30 | 60" | NS, ortho |
| A001 | M | 20/20 | 20/40 | >800" | 12Δ RXT |
| A002 | M | 20/20 | 20/70 | >800" | 16Δ RXT |
| A004 | M | 20/20 | 20/40 | >800" | 10Δ LET |
| A005 | S | 20/20 | 20/60 | >800" | 8-10Δ RET |
| A006 | A | 20/20 | 20/70 | >800" | NS, ortho |
| A007 | A | 20/20 | 20/30 | >800" | NS, 6Δ XP |
| A009 | A | 20/20 | 20/30 | 100" | NS, 8-10Δ XP |
| A010 | M | 20/25 | 20/60 | >800" | Constant alternating 10Δ ET |

Participants viewed drifting dichoptic plaids consisting of orthogonal gratings (1 cpd, 4 deg diameter, 0.25 cycles/s, 100% contrast, 60 seconds per trial, 42 trials) through a mirror stereoscope (Figure 1a). Alignment was achieved using a Nonius cross. Through the stereoscope, the dominant eye always viewed a grating oriented 45° moving up and to the left, and the non-dominant eye always viewed a grating oriented 135° moving up and to the right. As the gratings underwent binocular rivalry, participants continuously reported their form percept using 3 keys (single grating oriented 45°, single grating oriented 135°, or any partial or complete piecemeal combination). Participants were instructed that any mixture of the two gratings, even partially, should be considered as piecemeal. Motion direction percepts were reported by using a mouse to control an on-screen arrow. Participants were given an unlimited practice period to master the controls prior to starting the experiment. To ensure participants were responding accurately, 6 catch trials with monocularly presented stimuli were randomly interleaved within trial blocks.

**Figure 1.**
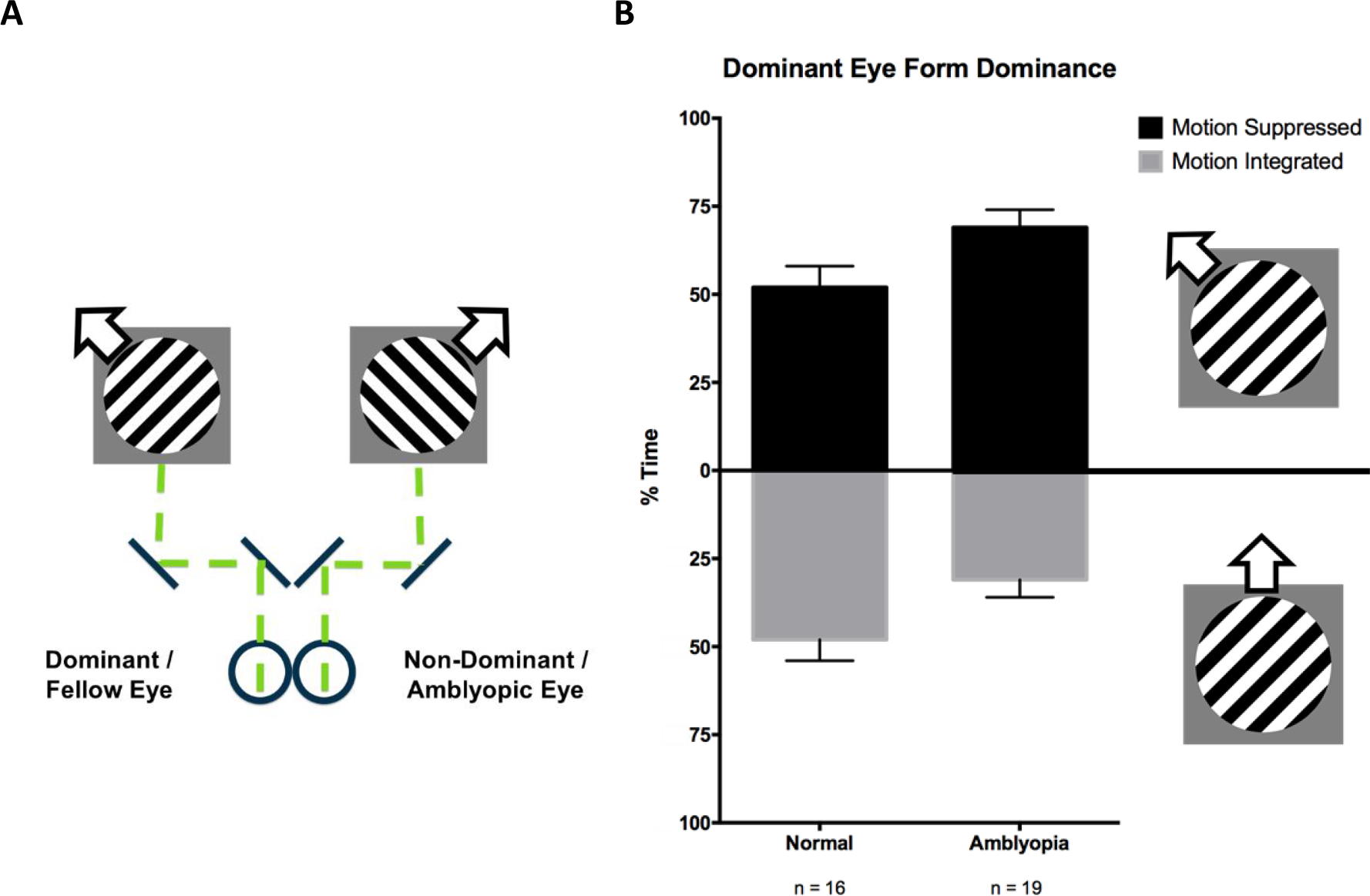
(A) Dichoptic presentation of plaid stimuli using two orthogonal drifting gratings. Arrows indicate direction of motion. (B) Extent to which binocular integration of motion information occurs during periods of non-dominant/amblyopic eye suppression. Figure designed in style as per Andrews & Blakemore(8).

Responses were binned based on different combinations of form and motion responses into 8 categories: complete dominant eye dominance, complete non-dominant eye dominance, binocular form and motion integration, transparent motion, form suppression of the dominant or non-dominant eye (with concurrent motion integration) and motion suppression of the dominant or non-dominant eye (with concurrent form integration). When only the dominant eye grating was visible, motion information was categorized as either suppressed (polar angle of motion direction arrow between 113–158°) or integrated (polar angle of motion direction arrow between 68–112°). Time spent in each motion direction category was represented as a proportion of the total time for which only the dominant eye grating was visible. An independent samples t-test was used to compare the proportion of time spent in each motion direction category between the control and amblyopia groups.

## Data Availability

Data and analysis code are available online in the University of Waterloo's repository.

## Acknowledgments

Supported by a NSERC PGS-D Grant (AC) and a NSERC Discovery Grant (BT; Ottawa, ON, Canada)

## Author Contributions

A.C., B.T. designed research; A.C., K.T., G.N., C.H. performed research; A.C., A.S., B.T. analyzed data; A.C., A.S., C.H., B.T. wrote the paper.

